# The ratio of interacting miRNAs’ expressions is a robust biomarker for disease classification in multi-center data

**DOI:** 10.1101/2023.06.29.23291976

**Authors:** Yonghao Zhang, Cuidie Ma, Rui Ding, Han Chen, Lida Xu, Changyuan Yu

**Author notes:** Corresponding authors: Lida Xu,; Changyuan Yu. These authors contributed equally to this work (Yonghao Zhang, Cuidie Ma).

## Abstract

**Background:** Many miRNA-based diagnostic models have been constructed to distinguish diseased individuals. However, due to the inherent differences across different platforms or within multi-center data, the models usually fail in the generalization for medical application.

**Results:** Here, we proposed to use the within-sample expression ratios of related miRNA pairs as markers, by utilizing the internal miRNA: miRNA interactions. The ratio of the expression values between each miRNA pair turned out to be more stable cross multiple data source. Moreover, we adopted the genetic algorithm to solve the curse of dimensions when exploring the features.

**Conclusions:** The application results on three example datasets demonstrated that the expression ratio of interacting miRNA pair is a promising type of biomarker, which is insensitive to batch effects and has better performance in disease classifications.

## Introductions

MicroRNAs (miRNAs) have emerged as valuable biomarkers for the early diagnosis of diseases due to their tissue-specific expression profiles and better specificity [1]. However, the expression levels of miRNAs may vary across different platforms or protocols, which limits the application of diagnostic models. This phenomenon, known as batch variance, is prevalent in all types of high-throughput biological platforms[2], and exists commonly in multi-center data[3–6]. The difference in data distribution from multiple centers is an obstacle to obtaining reliable conclusions in joint analysis, and it prevents the models learned in one dataset from working in other external datasets[3,7,8]. Thus, effectively handling batch effects in the integration of different datasets is one of the frontiers in large-scale biological data analysis[9].

Several batch effect correction methods have been developed to facilitate the joint use of multi-center data. For example, the ‘ComBat-seq’ tool based on the negative binomial regression model was developed specifically for RNA-seq count data[10]; the ‘removeBatchEffect’ function in ‘limma’ package can be used to correct the data variation caused by the batch effects[11]. However, these correction methods force the data shapes to be transformed artificially, which may introduce false discoveries[12] In contrast, the intrinsic regulatory pathways are not affected by experimental conditions, which makes the relationships between genes have the potential to be a type of normalizer-free and batch-insensitive markers. Under this consideration, we propose the ratio of the expression values between related miRNAs (ERRmiR) as a promising novel form of biomarkers for facilitating aggregation analysis of data from multiple sources.

To discover ERRmiR features with biological significance, a miRNA interaction network is needed as prior knowledge. It is widely known that miRNAs not only regulate the expression of mRNAs but also target non-coding RNAs, including long non-coding RNAs and miRNAs[13]. miRNAs can directly bind to the 3’UTR of transcription factors (TF), which can also reversely activate or repress miRNA expressions [14]. For example, miR-181b affects the expression of miR-21 through the transcription factor FOS, a critical signaling protein for glioma progression[15]; miR-660-5p has been reported to control the expression of miR-486-5p via mouse double minute2 (MDM2) and p53 (also known as TP53) in a study of lung cancer[16]. A recent review also summarizes numerous examples of miRNA-TF and TF-miRNA interactions in various cancers, demonstrating the importance of the interaction between miRNA and pluripotent transcription factor in determining the occurrence of human cancers[14]. All these examples provide important clues for understanding the role of the TF-mediated miRNA functional network in tumor regulation.

In this study, we constructed a TF-mediated miRNA interaction network using public databases and demonstrated that the ERRmiR features were relatively insensitive to batch effects in multi-center studies. We then adopted a genetic algorithm in the feature screening process to avoid the dimension curse, which had a great capacity for selecting markers with stable performances in developing diagnostic models. Lastly, we used three independent examples involving plasma and tissue samples to illustrate this method and exhibit its effects.

## Materials and methods

### Construction of miRNA interaction network

The TF-mediated miRNA: miRNA interaction network was constructed by combining the data of miRNA-TF and TF-miRNA relationships. If a transcription factor regulated by miRNA_a was able to regulate miRNA_b, miRNA_a was assumed to be able to influence miRNA_b. Then, they were connected in the miRNA interaction network.

The data of those relationships were collected from several public databases. The microRNA-target interactions validated experimentally were collected from miRTarBase [17], among which 8014 targets were recognized as transcription factors based on the hTFtarget[18] and AnimalTFDB[19] databases. On the other hand, 1266 records of transcription factors regulating precursor miRNAs were obtained from the TransmiR[20] v2.0 database. Combining these two parts of data, a total of 51,770 miRNA:pre-miRNA indirect interactions were obtained. Then pre-miRNAs were mapped to mature miRNAs according to the mirbase gff3 file. Finally, the miRNA: miRNA interaction network included 75,507 unique records of the indirect interaction relationships.

### Feature generation

Features were generated by calculating the ratio of expression values between each related miRNA pair in the miRNA interaction network constructed above. miRNAs were filtered based on an expression threshold of 100 to ensure that miRNAs could be detected stably. To avoid the divisor being zero, the denominator was added by one. The feature constructed with the connected pair of miRNA_a and miRNA_b was denoted as ERRmiR(a,b), then the formula was as follows:

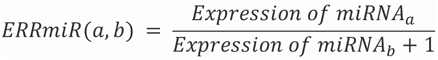

### Data collection and pre-processing

The data used to display the robustness of ERRmiR features on multi-source data from different library preparation kits were obtained from GSE133719 and GSE141658 datasets on Gene Expression Omnibus (GEO)[21] database. Three examples, including SARS-CoV-2-19, RCC, and LUAD projects, were used to verify the method in this study. Data of the three projects were collected from the NCI Genomic Data Commons (GDC)[22] database and GEO (detailed in Table 1). The miRNA expression matrices in the CPTAC[23]/TCGA[24] database were downloaded using the GDC tool. ExceRpt[25] was used to perform annotation and quantification of the raw data from GEO to obtain the expression matrices of miRNAs. For comparison of the results among different datasets within the same project, counts of reads were uniformly converted to RPM (reads per million mapped reads) values. In the SARS-CoV-2 project, the plasma of persons with non-severe symptoms (mild patients and healthy) were used as the controls, and the plasma of those with serious symptoms were used as the disease samples. In the RCC and LUAD projects, normal tissues were used as the controls, and primary tumor tissues were used as the disease samples.

**Table 1:**
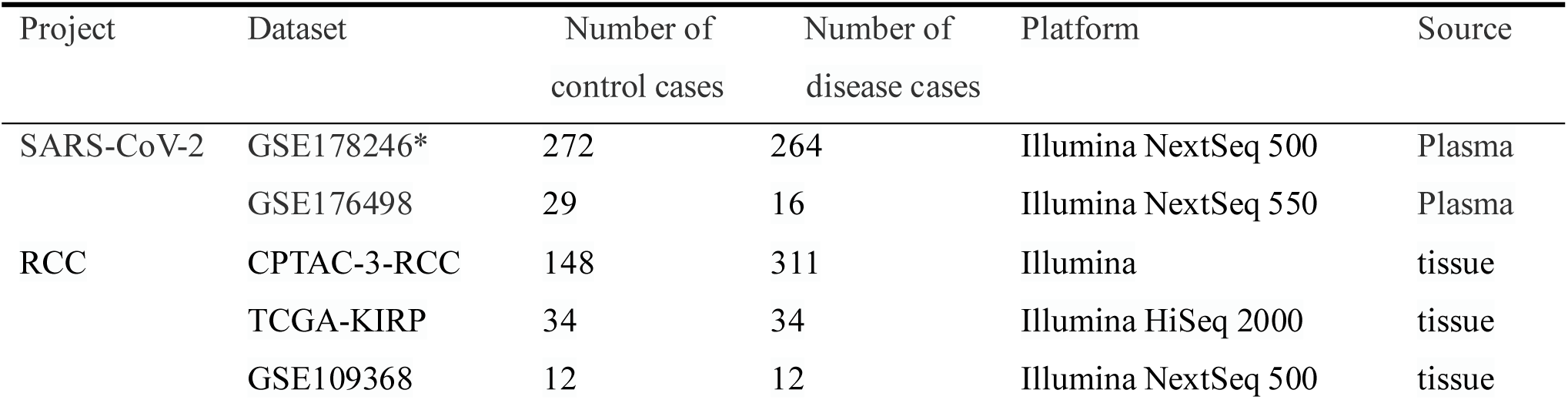
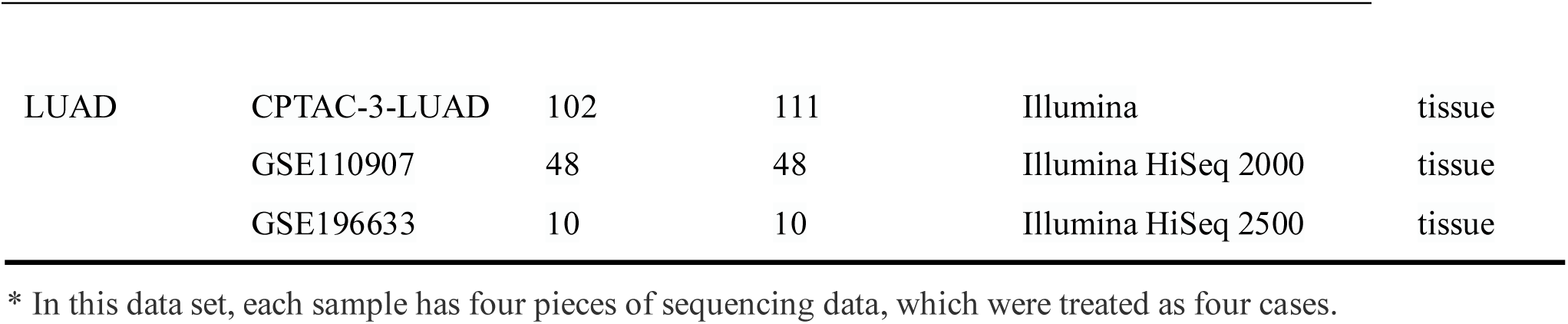
Sample information in detail

| Project | Dataset | Number of<br>control cases | Number of<br>disease cases | Platform | Source |
| --- | --- | --- | --- | --- | --- |
| SARS-CoV-2 | GSE178246* | 272 | 264 | Illumina NextSeq 500 | Plasma |
|  | GSE176498 | 29 | 16 | Illumina NextSeq 550 | Plasma |
| RCC | CPTAC-3-RCC | 148 | 311 | Illumina | tissue |
|  | TCGA-KIRP | 34 | 34 | Illumina HiSeq 2000 | tissue |
|  | GSE109368 | 12 | 12 | Illumina NextSeq 500 | tissue |
| LUAD | CPTAC-3-LUAD | 102 | 111 | Illumina | tissue |
|  | GSE110907 | 48 | 48 | Illumina HiSeq 2000 | tissue |
|  | GSE196633 | 10 | 10 | Illumina HiSeq 2500 | tissue |

\* In this data set, each sample has four pieces of sequencing data, which were treated as four cases.

### Feature screening and classification modelling

In each project, the dataset with the most samples was divided into a training set and a test set proportionally and randomly according to 0.75:0.25, and the training set was used to perform target screening. Univariate screening of the ERRmiR features was performed based on the foldchange of the mean expression in diseased samples compared to that in the controls and the p-adjust value of t-test between the two groups. The ‘sklearn-genetic’ package was used parallel for 100 times to obtain the optimal subsets of features. The features with higher appearance frequencies in the optimal subsets were selected as targets for the disease.

The ‘scikit-learn’ package was used to build models for disease classifications. During model training, the learning curves were used to detect whether the estimator was overfitting. The trained model was validated on a test set and other external validation datasets within the same project.

### Statistical analysis and visualization

The quartile plots of miRNA expression / ERRmiR feature values were drawn by the ‘matplotlib’ tool. The significance analyses were conducted using ‘scipy’. The miRNA network was visualized using ‘pyvis’ and ‘seaborn’ tools. In miRNA pathway enrichment analyses, target genes of miRNAs were first identified through the database ‘tarbase’ using the ‘multMiR’ package in R language, then pathway enrichments were performed using ‘clusterProfiler’.

## Results

### The schematic of ERRmiR signatures generation and screening

We developed a screening process for ERRmiR signature generation based on machine learning methods (Figure 1). We first constructed a miRNA interaction network by integrating several databases, including miRTarBase, hTFtarget, AnimalTFDB, and TransmiR v2.0. The network contained 75,507 unique records of indirect interaction relationships between miRNAs. We then calculated the expression ratios of related miRNA pairs as ERRmiR features. The screening dataset was randomly divided into a training set and a test set, and the features were filtered in the training set using univariate analyses such as t-test and the foldchange of the mean expressions between two groups. We used a genetic algorithm to screen the features, and those with higher frequencies in the screening processes were selected as candidate markers. The trained model was validated on the test set within the same screening dataset and evaluated on external validation datasets. This approach was suitable for discovering biomarkers for various samples.

**Figure 1:**
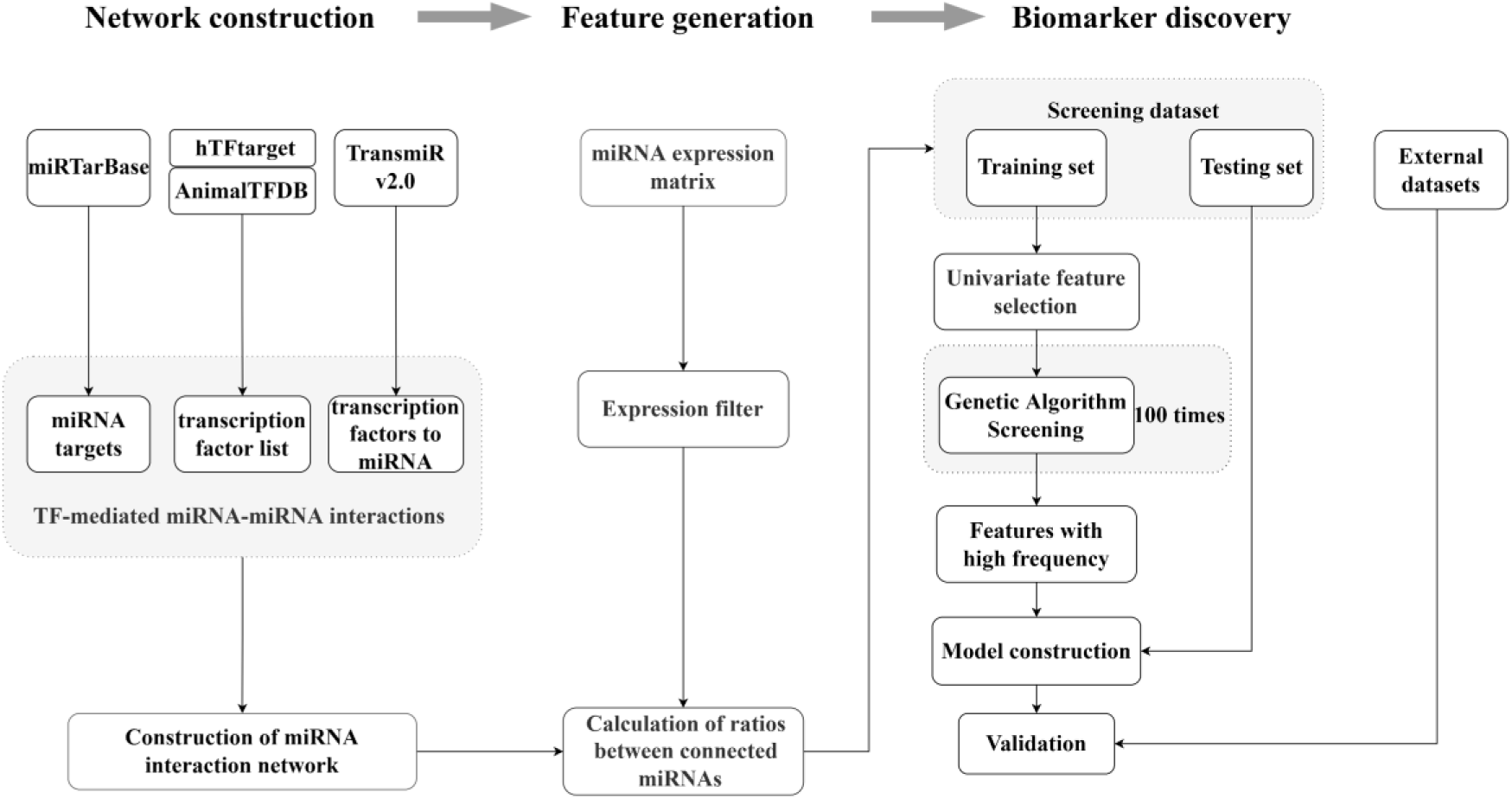
Overview of the ERRmiR marker discovery process. First, the miRNA network was constructed based on the TF-mediated interactions. Then, the ERRmiR features were calculated between the connected genes in the network as new variables for subsequent process. Finally, target screening and model construction were performed on the screening dataset and verified on the validation dataset.

### Construction of a miRNA interaction network

We constructed a miRNA interaction network based on indirect interactions mediated by transcription factors. The interactions mediated by transcription factors, induce the expression of one miRNA to impact the activation or inhibition of other miRNAs. Take miR-183-5p as an example to show how miRNAs regulate other miRNAs through transcription factors (**Figure 2A**). Here the pentagram-labeled miR-183-5p is a regulatory miRNA, which regulates the square-labeled transcription factors and further affects the round-labeled target miRNAs. The blue linkages represented the interaction of miR-183-5p acting on the transcription factors, and the pink linkages represented the effects of transcription factors on other miRNAs. The network contained 75,507 unique records of indirect interaction relationships between miRNAs. The miRNA interaction network was visualized, and its degree distribution and several topological characteristics were presented in **Figure 2B** and **Figure 2C**.

**Figure 2:**
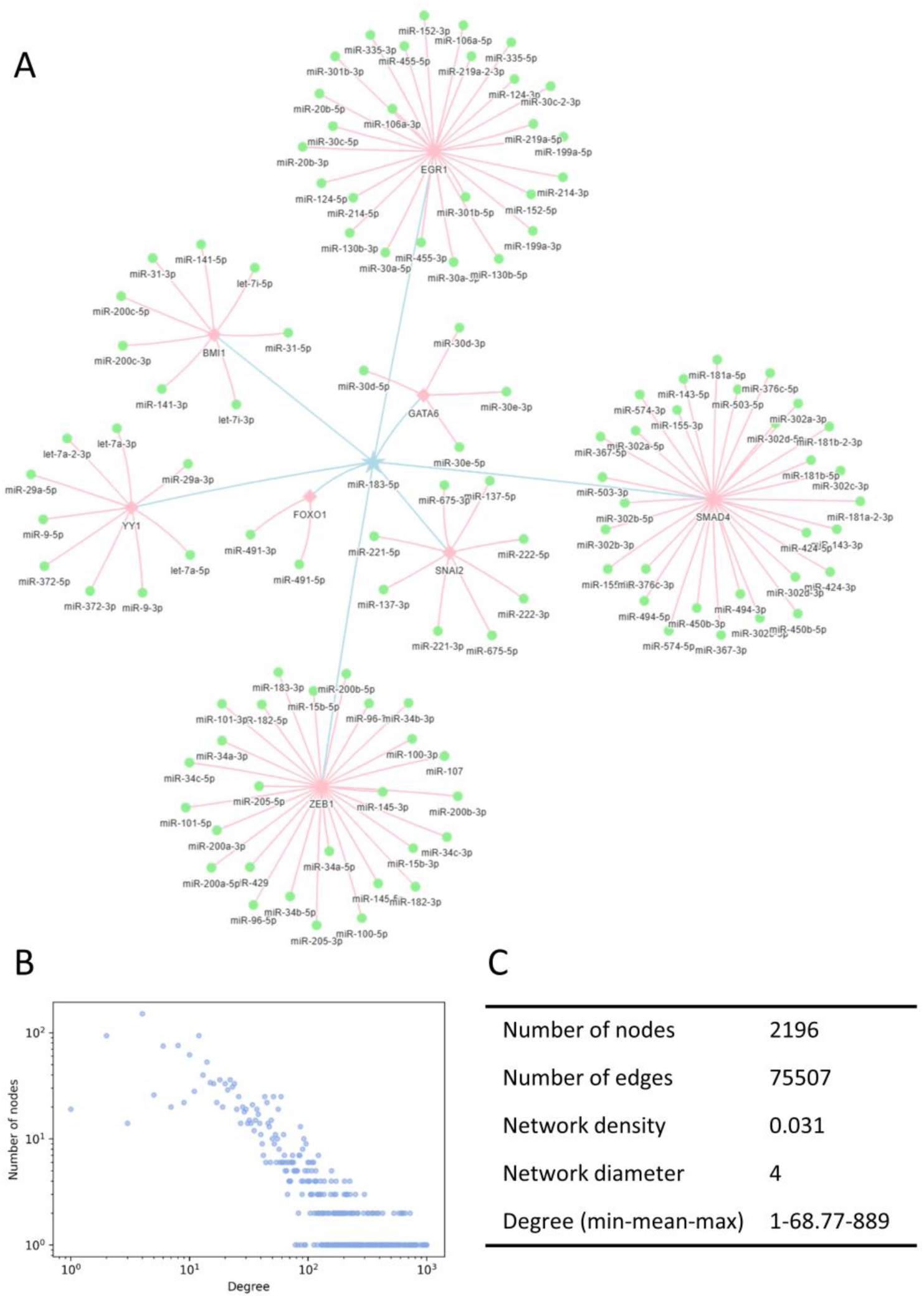
Illustration of the miRNA interaction network. (A) TF-mediated miRNA: miRNA indirect interactions. Pentagrams denoted the regulating miRNAs, squares denoted the transcription factors, and rounds denoted the regulated miRNAs. (B) Degree distribution of the miRNA interaction network followed a power-law tail. (C) Topological characteristics of the interaction network.

### Characterization of ERRmiR signatures

To verify the hypothesis that the expression ratios between interacted miRNAs would be more stable across multi-center data, the distribution of ERRmiR values was displayed compared to the distribution of the miRNA expression levels of the same samples (**Figure 3**). The experiment was about the sequencing data of the peripheral blood CD8+ T cells in triplicate from rheumatoid arthritis (RA) patients and healthy controls, by parallel receiving different library construction methods. The Quartile plots showed that the original miRNA expression data generated by different library preparation kits had significant differences on the scale and distributions (**Figure 3**, upper panel), while the distribution variation of ERRmiR values decreased (**Figure 3**, lower panel), which demonstrated the potential of ERRmiR features as batch-insensitive markers. We presented three application examples from various sample types and diseases.

**Figure 3:**
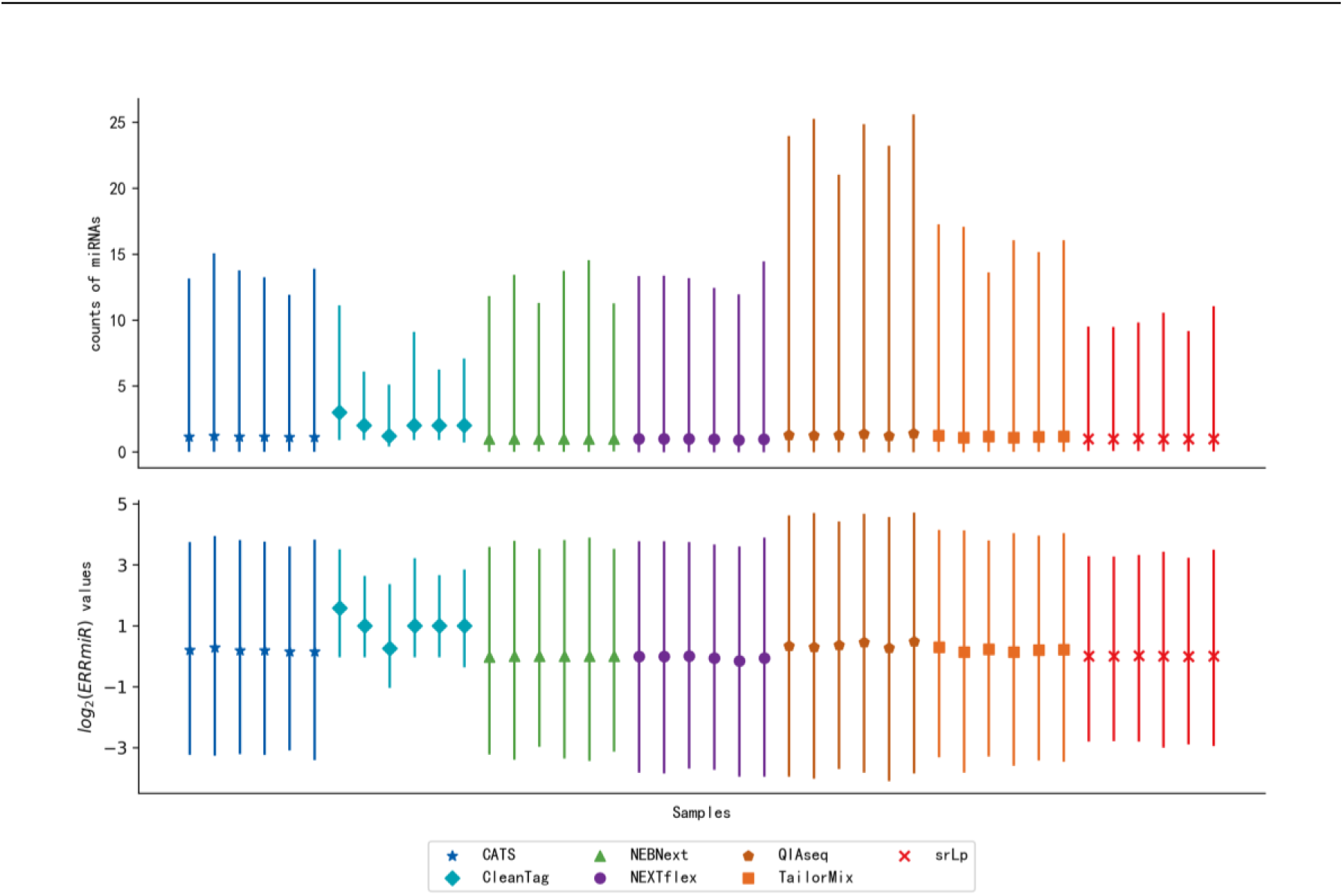
Quartile plots of miRNA expression (upper panel) and log2-ratios of every two miRNAs (below panel) for each sample. Each plot was represented with the median (a solid point), the 0.25 quartile and the 0.75 quartile of the distribution.

### Example 1: Classification of COVID-19 patients with severe symptoms using plasma samples

The advantage of ERRmiR features was first compared to the miRNA expression values on the dataset of COVID-19 plasma samples. The dataset GSE178246 was divided into a training set and a test set randomly, and the dataset GSE176498 was used as the external independent validation set. According to the protocol, there were 42 ERRmiR targets in total obtained during conducting the genetic algorithm for 100 times on the screening dataset. As shown in **Figure 4A**, the frequency distribution of target appearance was very steep: the highest frequency was up to 60, but there were only three targets with frequencies greater than 10. We selected the top 3 high-frequency features as markers, and tested them on the validation set. As expected, they were significantly different between the serious and non-serious groups (P<0.05) and showed relatively consistent trends across multiple datasets (**Figure 4B**). Based on these markers, an SVC model that was established on the training set, showed stable high performances on both the test set and the validation dataset (**Figure 4D**). To confirm the batch-insensitive nature of the ERRmiR features, the protocol of biomarker selection was also used on the expression matrix of miRNAs directly. As displayed in **Figure 4C**, the targets screened from the expression matrix of miRNAs lost effectiveness across batches of data, with miR-1224- 5p even showing opposite regulation trends. Accordingly, the model with miRNA expression values had a high AUC of 0.906 on the test set, but failed on the independent validation set with an AUC of 0.783 (**Figure 4E**). In addition, the five miRNAs that comprised the three ERRmiR markers were used for pathway enrichment, and the top 20 pathways were shown in Figure 4F. Infection pathways of bacteria and viruses, including Salmonella infection and Human papillomavirus infection, were significantly enriched.

**Figure 4:**
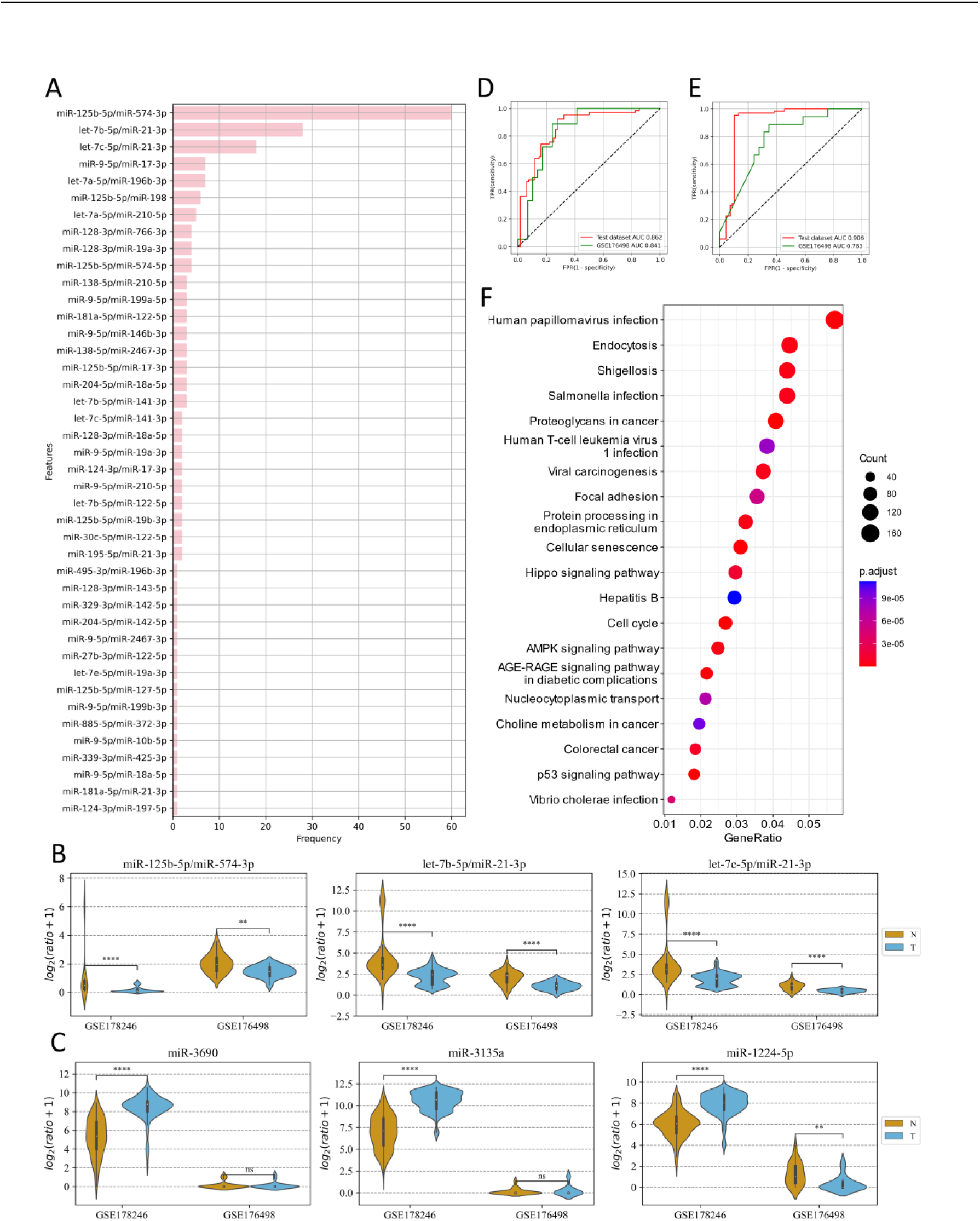
Analysis of ERRmiR features in the SARS-CoV-2 project. (A) The occurrence frequencies of the ERRmiR features in the 100 times genetic algorithm. (B-C) the top 3 high-frequency ERRmiRs showed relatively stable regulatory trend in both datasets rather than miRNAs. ROC curves of the models based on ERRmiR markers (D) and miRNA markers (E). (F) Pathway enrichment analysis of miRNAs involved in ERRmiR markers.

### Example 2: Diagnostic model of RCC using tissue samples

The strategy of marker discovery was also validated on the dataset of RCC tissue samples. Data from CPTAC-RCC dataset were used for screening targets and building the model. The TCGA-KIRP and GSE109368 datasets were used for external validations. After conducting the genetic algorithm, we obtained 115 targets with the frequency distribution shown in **Figure 5A**. As the same as in Example 1, the top 3 high-frequency ERRmiR features were selected as biomarkers, and presented significant differences between the cancer and control groups (P<0.05) with consistent regulation trends across multiple datasets (**Figure 5B**). Although part of the miRNAs in ERRmiR markers, such as miR-221-3p and miR-221-5p, didn’t display significant differential between the two sample groups in all the datasets (**Figure 5C**). A prediction model using the SVC algorithm was established on the screening dataset, and was able to achieve high AUC values on both two independent validation datasets (**Figure 5D**). The five miRNAs comprising the three ERRmiR markers were significantly enriched in several pathways associated with cancers (**Figure 5E**). Especially the p53 signaling pathway, and Hippo signaling pathway had been widely reported to be associated with RCC[26,27].

**Figure 5:**
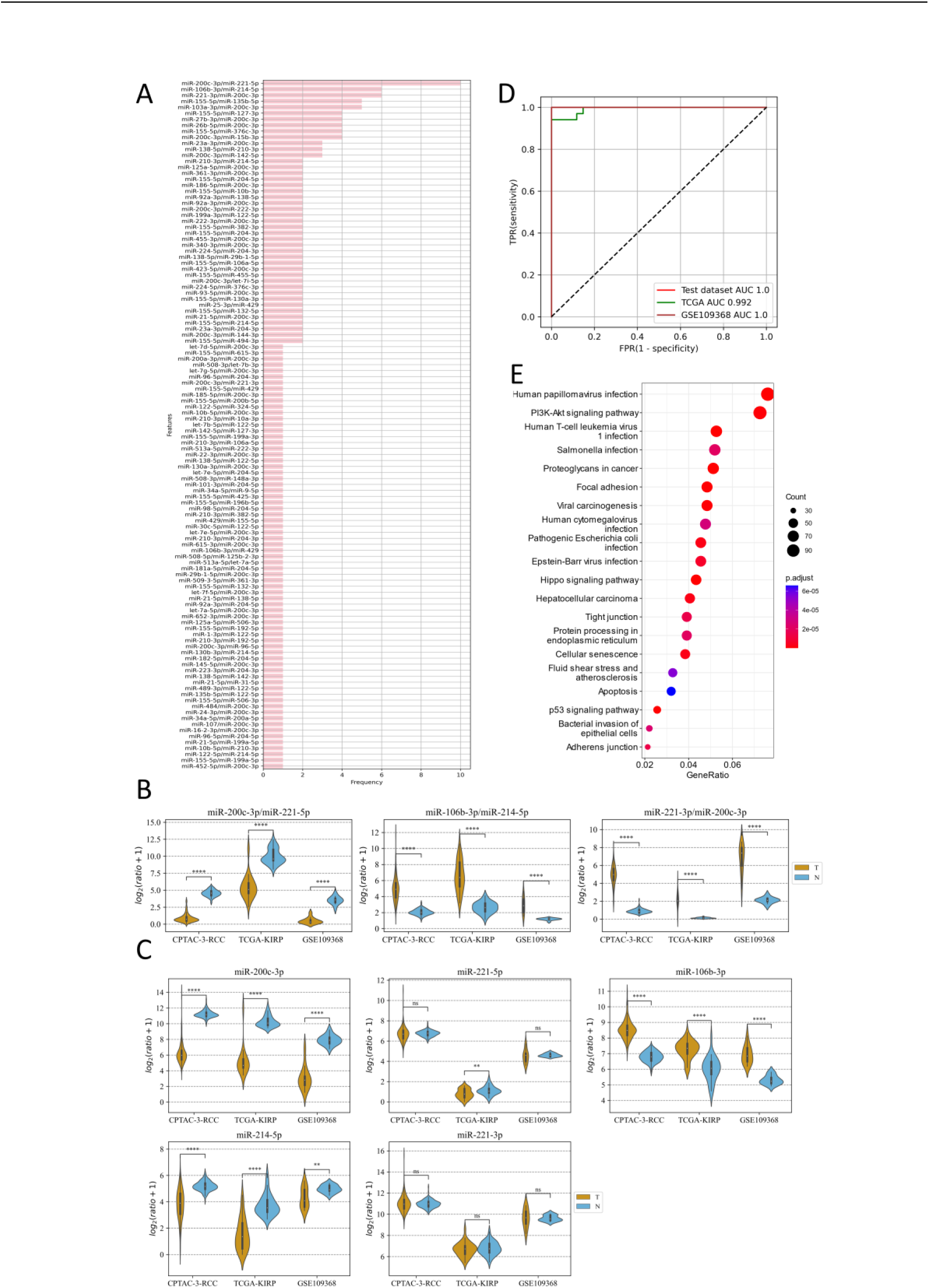
ERRmiR markers discovered in the RCC project. (A) Statistics of the frequencies of the ERRmiR features. Violin plots of the top 3 high-frequency ERRmiR features (B) and the miRNAs involved in them (C) among the three independent datasets. (D) ROC curves of the model based on the ERRmiR markers. (E) Pathway enrichment analysis of miRNAs in the ERRmiR markers.

### Example 3: Diagnostic model of LUAD using tissue samples

In the LUAD project, the CPTAC-LUAD dataset was used for screening targets and building the model. The GSE110907 and GSE196633 datasets were used for external validations. There were 31 targets obtained by conducting the genetic algorithm 100 times, with a relatively flat frequency distribution shown in **Figure 6A**. Then the top 3 high-frequency ERRmiR features were selected, and presented significantly differences between the cancer and control groups (P<0.05) with consistent regulation trends across multiple datasets (**Figure 6B**). The model trained in the screening set, had high AUC values of 0.995 and 0.91 in the GSE110907 and GSE196633 validation sets separately (**Figure 6C**). The five miRNAs comprising the three ERRmiR markers were significantly enriched in the p53 signaling, Cell cycle, PI3K-Akt pathways and so on, which had been widely reported to be associated with LUAD[28–30] (**Figure 6D**).

**Figure 6:**
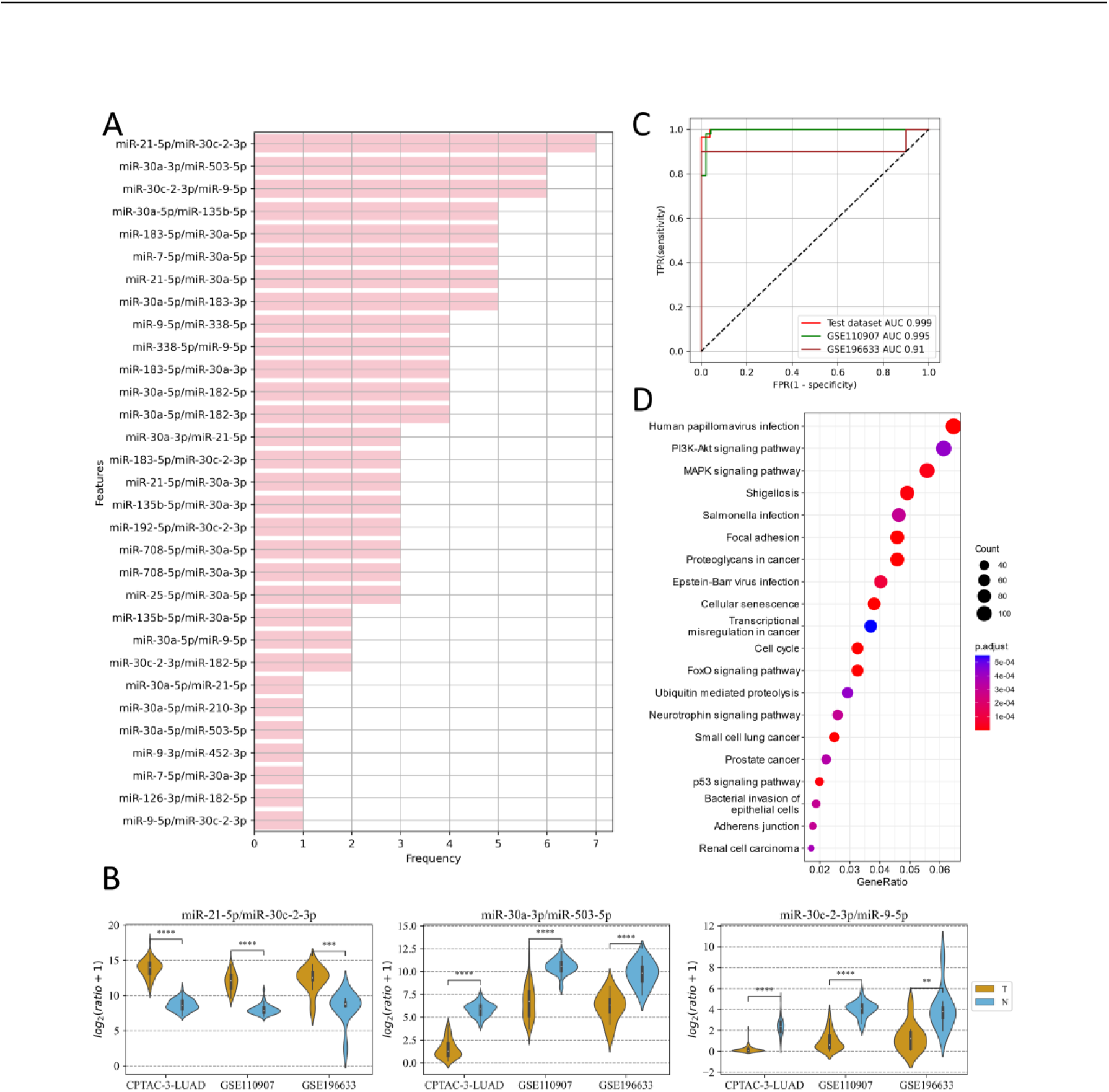
Discovery results on the LUAD project. (A) Statistics of the frequencies of ERRmiR features. (B) Violin plots of ERRmiR features ranked the top 3 by frequency. (C) ROC curves of the models based on ERRmiR markers. (D) Pathway enrichment analysis of miRNAs involved in the ERRmiR markers.

## Discussion and Conclusions

Using this protocol, we discovered some miRNAs with biological significance in all three examples, reflecting a low discovery rate in the ERRmiR markers. These miRNAs are disease-related and have been validated in previous studies. The results further demonstrate that the approach of this study is more helpful in implying the pathogenic mechanisms of diseases.

miRNA biomarkers have shown initial success in disease diagnosis and prognosis monitoring [31], but the noncontrollable experimental factors can cause deviation across different batches, making it difficult to use normalization of expression matrices alone for multi-center applications. In this study, we proposed an algorithm based on features formed by calculating the expression ratio of interacted miRNAs to remove batch effects. Coordinated with an integrated screening method utilizing the genetic algorithm, the algorithm can distinguish negative samples from positive samples on data from multi-sources. We demonstrated the effectiveness of this strategy at tissue and plasma levels with three examples, indicating its capacity for universal usage in developing diagnosis and classification models.

However, in previous studies, the lack of considering biological significance has led to improper strategies for construction and screening of expression ratio biomarkers. For example, some studies constructed expression ratio signatures by matching pairs with an upgraded gene and a downgrade gene, which ignores many worthy interactions[32,33]. Besides, this DE-dependent method would ignore many worthy interactions, and the construction method by pairing every two genes makes the number of features explode extremely, increasing the false discovery rate of targets and bringing tremendous pressure and difficulty to feature screening on small-sample biological data.

To address these issues, we constructed the expression ratio features based on a prior knowledge of miRNA interactions, which not only reduces the dimension of features but also helps to discover true relation markers. We included three types of miRNA:miRNA interaction (direct interactions, indirect interactions, and global interactions) summarized in a previous review[34] and considered the indirect miRNA interactions mediated by transcription factors. We constructed a TF-mediated miRNA interaction network to guide the generation of ERRmiR features, and with new miRNA regulatory relationships being discovered, the interaction network will likely expand, and new markers may gradually be revealed.

An efficient screening strategy is crucial to obtain stable biomarkers with excellent performance, especially for high-dimensional data and small sample size. In this study, we demonstrated that the expression ratios of miRNA pairs were more stable relative to the expression of individual miRNAs, and we preliminarily excluded low-expressed miRNAs to reduce the false discover rate and the dimension in calculating the feature matrix. The screening process comprised univariate analyses and multivariate genetic algorithm, and we repeated the genetic algorithm one hundred times to obtain high-frequency features, which were considered to be reliable.

Using this protocol, we discovered some miRNAs with biological significance in all three examples, reflecting a low discovery rate in the ERRmiR markers. Let-7b-5p, which is in a selected marker for predicting severe COVID-19 in the first example, has been reported to play a role in regulating ACE2 and DPP4 receptors and be significantly downregulated in nasopharyngeal swabs of patients[35]. Meanwhile, miR-21-3p which is regulated by let-7b-5p, shows an upregulation trend in this project and is consistent with the previous experiments of mice infected with SARS-CoV-2[36]. miR-106b-3p and miR-214-5p in the ERRmiR marker that has been selected in the RCC project, are both found to be critical oncogenes in previous studies. The high expression of miR-106b-3p may be an important factor in predicting poor prognosis in RCC patients[37,38], and the overexpression of miR-214-5p attenuates cell proliferation and metastasis[39]. In the LUAD project, the pairs containing miR-30a-3p or miR-30c-2-3p have been screened out. The role of the miR-30 family as tumor suppressors has been validated in previous reports[40], especially miR-30c-2-3p is reported to inhibit tumor progression in esophageal squamous cell carcinoma, breast cancer, and hepatocellular carcinoma[41–43]. miR-9-5p and miR-503-5p which are related with miR-30 in the markers, have also been reported to be associated with cell proliferation, migration, and invasion in non-small cell lung cancer[44,45]. These miRNAs are disease-related and have been validated in previous studies. The results further demonstrate that the approach of this study is more helpful in implying the pathogenic mechanisms of diseases.

## Data Availability

This data can be found here: https://portal.gdc.cancer.gov/ and https://www.ncbi.nlm.nih.gov/geo/.

https://www.ncbi.nlm.nih.gov/geo/

## Declarations

### Ethical approval and consent to participate

Ethical approval was not required for this study because we used a public database.

### Consent to publish

Not applicable.

### Availability of data and materials

Publicly available datasets were analyzed in this study. This data can be found here: https://portal.gdc.cancer.gov/ and http://www.ncbi.nlm.nih.gov/geo/. The dataset supporting the conclusions of this article is included within the article and its additional file. Source code is also available if required.

### Authors’ contributions

L.X. and C.Y. conceived and supervised the experiments. Y.Z., C.M., and L.X. wrote the manuscript. Y.Z., C.M., R.D. and H.C. performed the experiments. All the authors have read and approved the final manuscript.

## Acknowledgements

Not applicable.

## Additional file

**Additional file 1**

Table S1. A collection of ratio features generated based on transcription factor-mediated indirect action relationships of miRNAs.

Additional file 2

Table S2. Values of the features in Figure 4B.

Table S3. Values of the features in Figure 4C.

Table S4. Values of the features in Figure 5B.

Table S5. Values of the features in Figure 5C.

Table S6. Values of the features in Figure 6B.

